# sMAdCAM:IL-6 ratio influences disease progression and anti-viral responses in SARS-CoV-2 infection

**DOI:** 10.1101/2020.10.13.20182949

**Authors:** Dhanashree Jagtap, Vikrant M Bhor, Shilpa Bhowmick, Nandini Kasarpalkar, Pooja Sagvekar, Bhalchandra Kulkarni, Manish Pathak, Nirjhar Chatterjee, Pranam Dolas, Harsha Palav, Snehal Kaginkar, Sharad Bhagat, Itti Munshi, Swapneil Parikh, Sachee Agrawal, Chandrakant Pawar, Mala Kaneria, Smita D Mahale, Jayanthi Shastri, Vainav Patel

## Abstract

Recent studies positing the gut as a sanctuary site for viral persistence in SARS-CoV-2 infection highlight the importance of assimilating profiles of systemic as well as gut inflammatory mediators to understand the pathology of COVID-19. Also, the role of these markers in governing virus specific immunity following infection remains largely unexplored. A cohort (n=84) of SARS-C0V-2 infected individuals included a group of in-patients (n=60) at various stages of disease progression together with convalescent individuals (n=24) recruited between April and June 2020 from Mumbai, India. Follow-up of 35 in-patients at day 7 post diagnosis was carried out. Th1/Th2/Th17 cytokines along with soluble MAdCAM (sMAdCAM) levels in plasma were measured. Also, anti-viral humoral response as measured by rapid antibody test (IgG, IgM), Chemiluminescent Immunoassay (IgG) and antibodies binding to SARS-CoV-2 proteins were measured by Surface Plasmon Resonance (SPR) from plasma.IL-6 and sMAdCAM levels among in-patients inversely correlated with one another. When expressed as a novel integrated marker –sMIL index (sMAdCAM/IL-6 ratio), these levels were incrementally and significantly higher in various disease states with convalescents exhibiting the highest values. Importantly, sMAdCAM levels as well as sMIL index (fold change) correlated with peak association rates of receptor binding domain and fold change in binding to spike respectively as measured by SPR. Our results highlight key systemic and gut-associated parameters that need to be monitored and investigated further to optimally guide therapeutic and prophylactic interventions for COVID-19.

## Introduction

Having established itself in the human population, the SARS-CoV-2 virus and its associated pathology (COVID19) is a subject of intense study with implications for public health globallyz (1–3). Inflammation, especially that mediated by ‘cytokine storm’ seems to be the primary etiological factor driving progression of COVID-19 disease (4,5). Persistent viral infection in sanctuaries such as the gut together with post-infection immunological sequelae are also emerging as relevant disease management issues that need to be addressed (6–8). Recent studies have identified key inflammatory cytokines such as IL-6 and TNF that are dysregulated and appear to play a significant role in mediating pathology (9,10). Also, the role, if any, of these cytokines and the associated inflammatory response in governing virus specific immunity remains largely unexplored following both symptomatic and asymptomatic infection. Understanding and identifying such signatures may guide current therapeutic and vaccine development efforts to ensure optimal disease management for this emerging pathogen. In this study, we undertook to examine cross-sectionally and longitudinally, profiles of inflammation, including a gut associated marker governing mucosal lymphocyte migration, across various stages of disease progression following SARS-CoV-2 infection. Concurrent evaluation of virus specific humoral immunity was also carried out to understand the link between these profiles and generation, persistence and real-time binding kinetics of antibodies against various viral antigens. Our results highlight a putative immunological marker that seems to govern both disease progression as well as generation of anti-viral antibodies to the neutralizable receptor binding domain of SARS-CoV-2.

## Materials and Methods

### Study population, setting, and data collection

A total of 60 in-patients and 24 convalescent individuals were recruited (from April-July, 2020), following informed consent, for the study from the isolation ward at Kasturba Hospital for Infectious Diseases (Municipal Corporation of Greater Mumbai). Pregnant women, prisoners, and children (less than 18 years of age) were excluded from the study. The Kasturba Hospital and ICMR-NIRRH institutional ethics committees approved this study. We obtained demographic data, clinical history at presentation, and laboratory results during admission.

Blood samples for the study were handled in accordance with ICMR guidelines for biosafety. Whole blood (1-3 ml) was collected in EDTA vacutainers and plasma was separated by centrifugation at 425 g for 10 minutes. IgG and IgM antibodies against SARS-CoV-2 were detected in fresh plasma samples using Rapid test from either Voxpress (Voxtur Bio LTD, India) or Tell me fast (Biocan Diagnostics Inc., Canada) kits and also by ARCHITECT™ Abbott™ (Abbott Diagnostics, USA) chemiluminescence immunoassay (CLIA) directed against SARS -CoV-2 anti-NC IgG. Remaining plasma samples were aliquoted and stored at −80° C until batch analysis of cytokines, Soluble MAdCAM and antibody binding profile using Surface Plasmon Resonance.

### Cytokine Bead Array

Human Th1/Th2/Th17 cytokines kit (BD CBA kit Cat no. 560484, NJ, USA) was used to measure IL-2, IL-4, IL-6, IL-10, TNF, IFN-γ and IL-17 cytokine levels in plasma of study participants. Briefly, 50 µl of plasma sample and standard were added to 50 µl of capture bead mixture followed by addition of 50 µl of Human Th1/Th2/ Th17 PE Detection reagent. Following incubation at room temperature in dark for 3 hours, the assay tubes were washed with 1 ml of wash buffer. The tubes were centrifuged at 200 g for 5 min and bead pellet was re-suspended in 300 µl wash buffer and acquired using BD Accuri™ C6. Five thousand events were acquired gated on selected bead population and analysis was carried out using BD C6 Accuri analysis software. GraphPad Prism was used to extrapolate individual cytokine concentrations in each sample.

### MAdCAM ELISA

Human MAdCAM-1 DuoSet ELISA kit (R&D Systems-DY6056-05, Minneapolis, USA) was used for estimation of soluble MAdCAM-1 in plasma in accordance with manufacturer’s protocol. Briefly, wells coated with capture antibody and reagent diluent (1% BSA in PBS) was added for blocking. MAdCAM standard or 1:100 diluted plasma was added to wells and incubated overnight at 4°C. Incubation with detection antibody was followed by addition of streptavidin-HRP which was allowed to react with substrate solution to give chromogenic readout. Reaction was stopped using 2N H_2_SO_4_ and absorbance measured at 450 nm and 570 nm using Synergy H1 microplate reader (Biotek, Vermont, USA). Readings at 570 nm were subtracted from 450 nm and used to plot standard curve. Plasma concentration of sMAdCAM was calculated by interpolating absorbance values and multiplying by dilution factor.

### Surface Plasmon Resonance (SPR) studies: Binding of COVID-19 patients plasma to immobilized SARS-CoV-2 proteins-Spike, Receptor Binding Domain (RBD) and Nucleocapsid

The binding analysis of real time interaction between COVID-19 patients plasma and SARS-CoV-2 proteins was carried out using the SPR spectrometer (Biacore 3000, GE Healthcare Bio-Sciences AB, Uppsala, Sweden). Flow cells of CM5 (carboxmethylated dextran matrix) sensor chip were first activated with 0.2 M 1-ethyl-3-(3-dimethyl aminopropyl) carbodiimide hydrochloride and 0.05 M N-hydroxysuccinimide. SARS-CoV-2 proteins (Spike, RBD and Nucleocapsid) at a concentration of 20 μg/ml each in 10 mM sodium acetate, pH 4.5, was immobilized on the activated CM5 chip to about 1400 response units (RUs), and the unreacted groups were blocked with 1.0 M ethanolamine-HCl, pH 8.5. Flow cell, which was not immobilized with any protein, served as negative control.

Human COVID-19 patients plasma was divided into four groups based on their IgM/IgG status (Rapid Antibody Tests) and was used to determine the binding of antibodies to three SARS-CoV-2 proteins. Diluted plasma (1:10) in 10 mM HBS-EP, pH 7.4 running buffer was injected over Spike, RBD & Nucleocapsid immobilized CM5 chip for 150 seconds (association phase) followed by subsequent dissociation for 150 seconds and recording of their spectra. All the experiments were performed at 25 °C, and the flow rate used was 20 μl/min. The surface at the end of each experiment was regenerated using a 25 μl pulse of 10 mM Glycine-HCL, pH 2.5 to remove any bound analyte. Sensorgrams were corrected by subtraction of the signal from the negative control surface as well as pre COVID-19 plasma.

Data were evaluated using the BIAevaluation software program (4.1 version, GE Healthcare Bio-Sciences AB). The 1:1 Langmuir model was used for fitting the experimental data and to calculate the association response differences and dissociation rates (kd).

### Statistical analysis

Statistical analysis was performed in GraphPad Prism 8 using non parametric tests. Statistical significance of differences between groups were assessed using Mann-Whitney U-test. Follow up data from same individual was compared with Wilcoxon matched-paired signed rank test. Spearman’s rank-order correlation was used to analyse the association between participant attributes and statistical significance was accepted at p<0.05.

## Results

### Demographic and clinical characteristics of in-patients and convalescent individuals

During the period from April through July 2020, 60 in-patients with confirmed Covid-19 infection and under isolation were recruited at Kasturba Hospital. Of these 35 were followed up 7 days post admission. A total of 24 individuals, clinically documented to have been recovered and discharged were also recruited as convalescents. Convalescent individuals had 2 consecutive negative respiratory swabs or were cleared of symptoms prior to discharge and their blood samples were collected atleast 2 weeks post discharge. The demographic, serological and clinical characteristics of the patients are shown and summarized in Supplemental Table 1 and Table 1 respectively. As apparent from these tables our study cohort included individuals that were analysed in a cross-sectional, longitudinal (0 day and 7 day time points) manner and at clinically distinct (in patients and convalescent) stages of disease progression. Also, as expected, males pre-dominated and individuals with co-morbidities constituted a significant proportion (28/66; 42%) of the cohort. Serological data evaluated through both a rapid test (IgG, IgM) and CLIA (anti-NC IgG) showed an overall concordance of 77% for IgG and also allowed us to group these individuals based on IgG and/or IgM positivity. When grouped in this fashion it was interesting to note that while a majority of convalescent individuals ∼88% (21 out of 24) were seropositive for SARS-CoV-2 about 12% (3 out of 24) were seronegative both by CLIA and rapid test (Table 1 and Supplemental Table 1). Also, as a group, convalescent individuals seemed to have lower levels of circulating anti-viral antibodies following infection.

**Table 1.** Abridged clinical data of study participants

|  | **In-patients**<br>**(n=60)** | **Convalescent**<br>**(Group V)**<br>**(n=24)** |
| --- | --- | --- |
| Number of Participants | 60 (35 follow up<sup>b</sup>) | 24 |
| Age<sup>a</sup> (years) | 46 (18-74) | 34 (20-55) |
| Gender | 8 Women (22%)<br>28 Men (78%) | 3 Women (14%)<br>19 Men (86%) |
| Days post PCR test until<br>sampling<sup>a</sup> | 0-19 (6) | 19-62 (37) |
| CLIA index<sup>a</sup> | 1.55-9.69 (5.93) | 2.58-9.47 (4.48) |
| Grouping |  |  |
| I IgM-IgG- | 38 | 3 |
| II IgM+IgG- | 13 | - |
| III IgM+IgG+ | 25 | 2 |
| IV IgM-IgG+ | 17 | 19 |

**Footnote:** <sup>a</sup> Data are expressed as the median (range) <sup>b</sup> Blood sampling of 35 study participants was carried out on Day 0 and Day 7 and considered separately (as follow-up samples) for analysis. Sero-grouping data was not available for 1 in-patient (follow-up).

### Inflammatory cytokine profile and gut migration

As shown in (Supplemental Figure 1A and 1B), elevated levels of IL-6 and TNF were observed both in-patients as well as, of note, in convalescent individuals compared to healthy individuals. Also, these levels spanned a range of sero-positivity in terms of virus specific IgG and/or IgM. Also, we noticed a significant reduction of IL-6, but not TNF, as seropositivity transitioned from double negative to IgG, IgM double positive and finally to class switched (IgG single positive) responses in convalescent individuals. Increased levels of these cytokines did not correlate with initial Ct values, each other and that of any other cytokine measured (Supplemental Figures 2 and 3). When these levels were examined longitudinally (Supplemental Figure 4) in a group of 11 individuals that were followed at day 0 (day of diagnosis) and day 7 (post diagnosis) we noticed a declining trend for IL-6 but not TNF.

Additionally and for the first time we report on gut inflammatory/migratory marker sMAdCAM (Figure 1 and Supplemental Figure 1C) which appeared to be circulating at levels lower than those seen in healthy individuals and convalescents. Also there was no difference between sMAdCAM levels in 34 individuals that were followed at day 0 and day 7. Interestingly however (Supplemental Figure1c), in contrast and conversely with respect to IL-6, sMAdCAM levels seemed to reduce (compared to healthy controls) following early infection (IgM single positive and IgG, IgM double positive stages) and attain restoration in class switched individuals (IgG single positive and convalescents). Further, LPS levels, evaluated in a subset of in-patients and convalescents, did not vary signifying absence of microbial translocation associated gut inflammation (Supplemental Figure 5).

**Figure 1:**
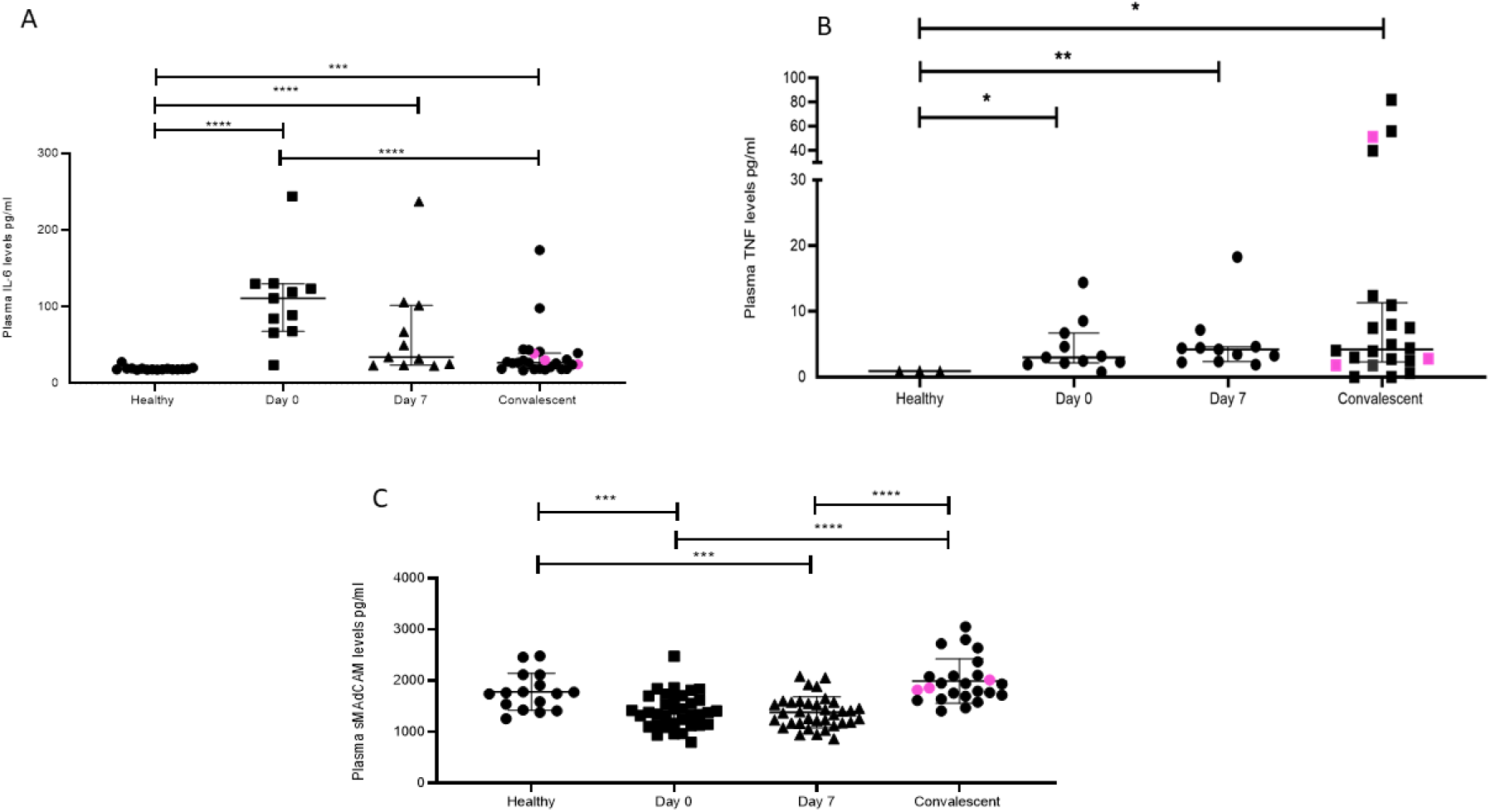
Dynamic changes in plasma inflammatory markers of SARS-CoV-2 infected individuals and convalescent individuals. Variation in levels of (A) IL-6 at day 0 (n=11), day 7 (n=11), in convalescent (n=22) and in healthy (n=19) group (B) TNF at day 0 (n=11), day 7 (n=11), in convalescent (n=22) and in healthy (n=3) group and (C) Soluble MAdCAM at day 0 (n=35), day 7 (n=35), in convalescent (n=24) and in healthy (n=19) group. Pink coloured symbols indicate IgG-/IgM-individuals in convalescent group. Statistical significance was calculated by Wilcoxon matched-pairs signed rank test and Mann-Whitney U-test; *, p < 0.05; **, p < 0.01; and ***, p<0.001.

Next, analysing these individuals based on disease progression clearly demonstrated a declining circulating IL-6 but not TNF concentration (Figure 1). Stratification of the in-patient data (Supplemental Figure 6) on the basis of gender revealed a marked difference in circulating levels of IL-6 and sMAdCAM where women had significantly lower and higher levels of these respectively.

### sMAdCAM:IL-6 ratio (sMIL index): A novel biomarker for disease progression

Results obtained from the heretofore described analyses (Figure 1 and Supplemental Figure 6) suggested that sMAdCAM and IL-6 levels independently seemed to track (Supplemental Figures 7) with disease progression, albeit in opposing directions. Suspecting an underlying association between both of these apparently unrelated signatures, we examined the utility of their being combined as an integrated marker that could better correlate and discriminate between various stages of infection.

Remarkably, when levels of both these analytes were expressed as a ratio we observed (Figure 2) a clear delineation of in-patient and convalescent data into discrete categories based on disease progression (Figure 2A and 2B), gender (Figure 2C), time from infection (Figure 2D) and intriguingly serological status (Figure 2E). These observations seemed to posit a role for this index in the development of antibodies against SARS-CoV-2.

**Figure 2:**
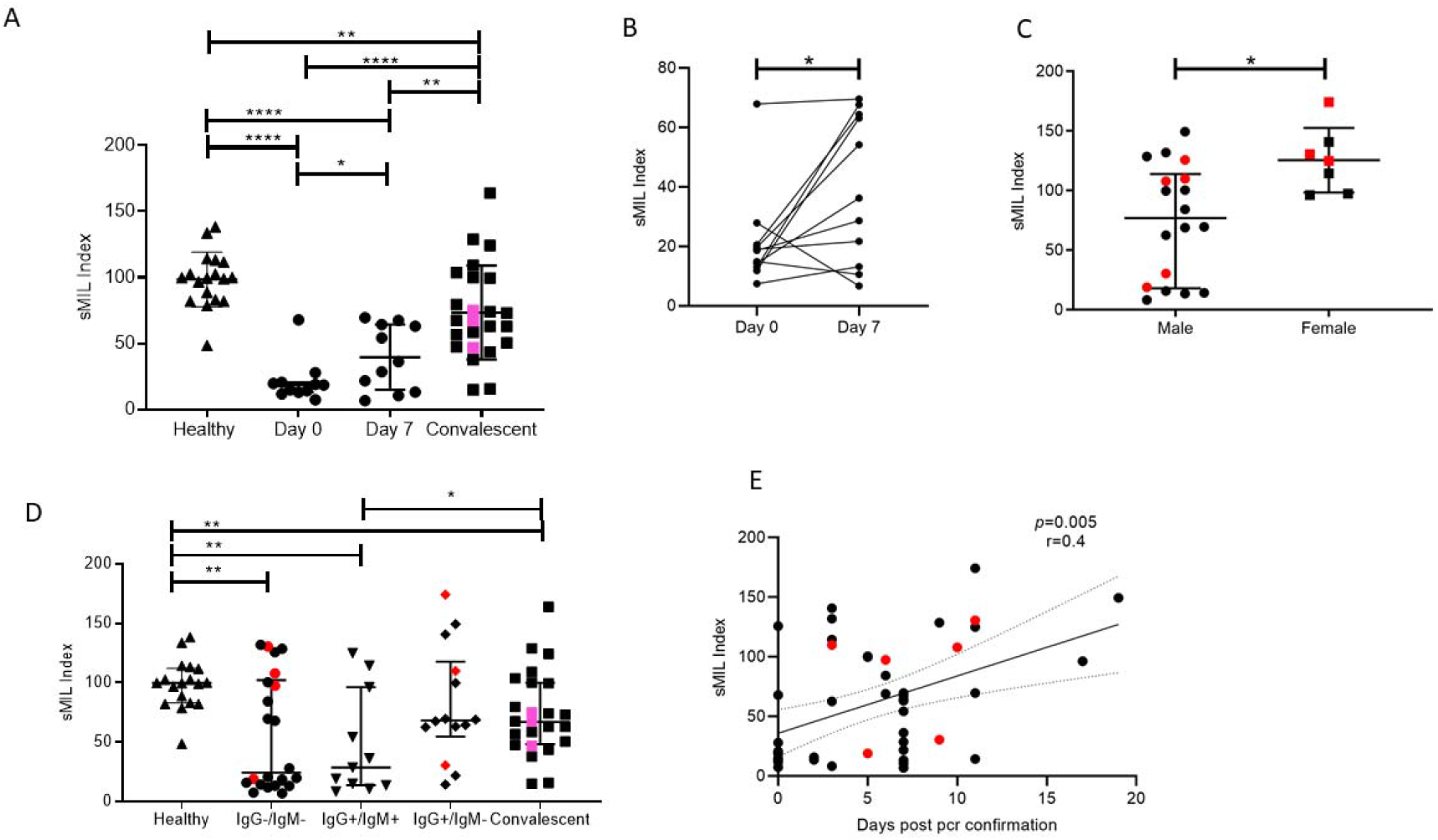
Analysis of soluble MAdCAM and IL-6 ratio (sMIL index) with SARS-CoV2 disease progression A. Variation in sMIL index at day 0 (n=11), day 7 (n=11), in convalescent (n=22) and in healthy (n=19) group. Pink coloured symbols indicate IgG-/IgM-individuals in convalescent group. B. Change in sMIL index between day 0 (n=11) and day 7 (n=11) expressed per individual. C. Differences in sMIL index between male (n=18) and female (n=7). D. Comparison of sMIL index between healthy (n=19),IgG-/IgM-(n=22), IgG+/IgM+ (n=11), IgG+/IgM-(n=14) and convalescent (n=22) groups.. E. Association of sMIL index with days since SARS-CoV-2 confirmation by PCR (n=47). Red coloured symbols represent asymptomatic individuals Statistical analysis was performed using Graphpad Prism 8.0 Wilcoxon matched-pairs signed rank test was used to compare paired samples of Day 0 and Day 7. Mann-Whitney U-test was used to compare unpaired groups. *, p < 0.05; **, p < 0.01, ***, p<0.001; and ****, p<0.001. Correlation analysis was performed using non parametric Spearman Rank Correlation test.

### Anti-viral responses and the role of inflammatory markers

Anti-SARS-CoV-2 responses were measured using surface plasmon resonance (SPR) that allowed us to obtain a sensitive and real-time evaluation of antibody binding to viral Spike, RBD and NC proteins. First, we demonstrated that SPR was indeed picking up antibody binding through the observed concordance between CLIA profile and viral protein binding association rates observed in our prospective cohort (Supplemental Figure 8). Further, we observed that individuals that were below the level of detection in the CLIA assay (Y-axis value 0) did have detectable binding using SPR.

We then proceeded to stratify plasma binding data (association rates) based on disease progression (Supplemental Figure 9) as well as IgM/IgG serostatus (Figure 3). In the case of the former we observed significant increases by day 7 for all 3 viral targets with increased responses against NC and RBD compared to S followed by steep declines of these responses in convalescent plasma.

**Figure 3:**
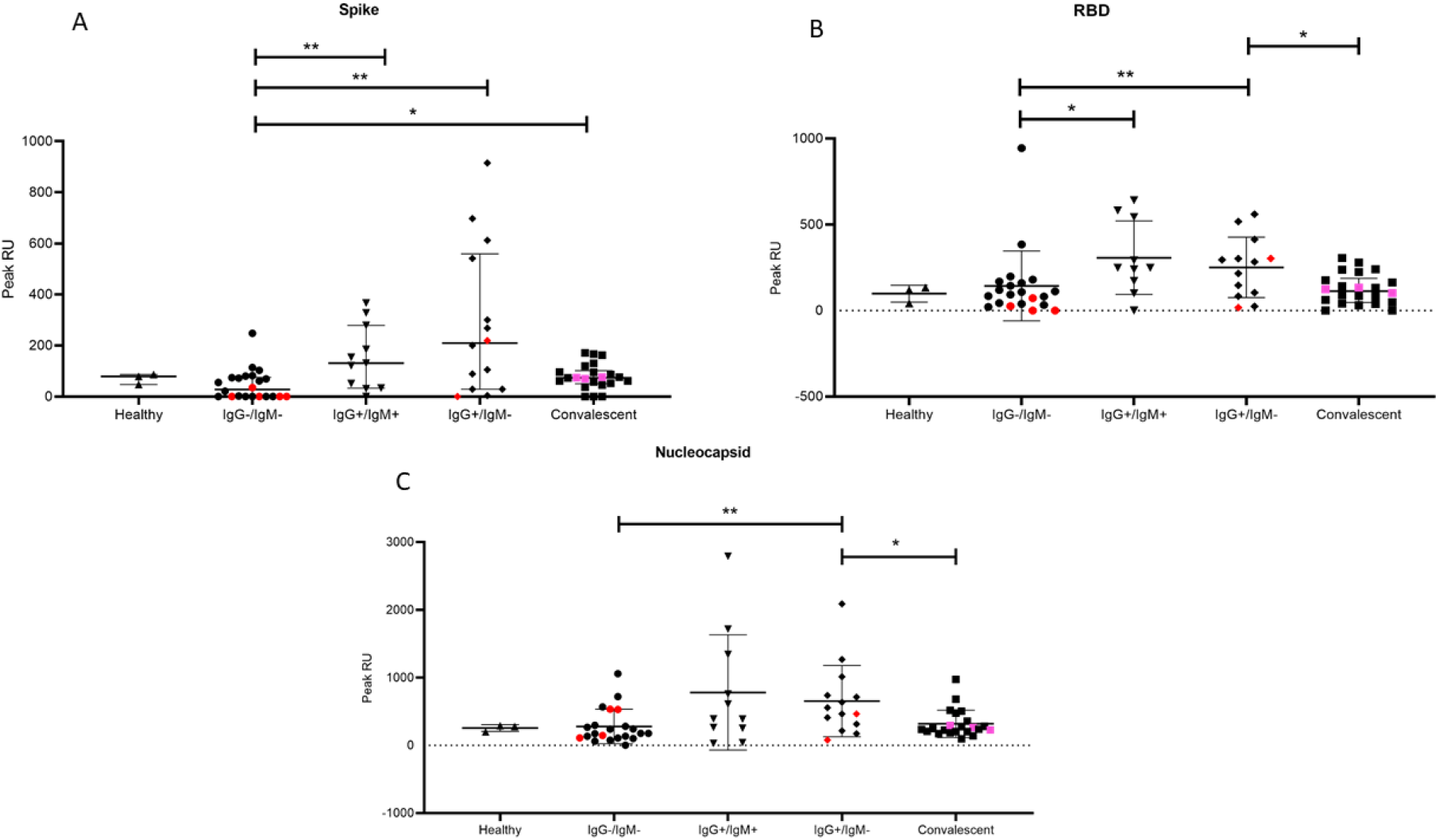
Distribution of Peak RU across various serological groups of SARS-CoV-2 infected individuals. Comparison of peak RU for (A) Spike(B) RBD and (C) Nucleocapsid between healthy (n=3), IgG-/IgM-(n=22), IgG+/IgM+ (n=11), IgG+/IgM-(n=14) and convalescent (n=22) groups. Red coloured symbols represent asymptomatic individuals. Pink coloured symbols indicate IgG-/IgM-individuals in convalescent group. Statistical significance was calculated by Mann-Whitney U-test; *, p < 0.05; **, p < 0.01; and ***, p<0.001

When grouped cross-sectionally by sero-status we observed (Figure 3), as would be expected, an increase in binding from the IgG-IgM-stage to the single positive, post class-switched stage (IgM-IgG+). Also, as observed in the smaller prospective cohort, association rates were higher for NC and RBD. For all three viral proteins, these rates were significantly lower in convalescent individuals with presumably mature memory B cells and viral clearance.

Having already demonstrated importance of circulating IL-6 and sMAdCAM either in combination (sMIL index, Figure 2) or in isolation (Figure 1 and Supplemental Figures 5 and 6) in influencing disease progression and anti-viral antibody production, we investigated the possibility of identifying a specific viral target (Spike, RBD or NC) whose binding response was associated and thus potentially attributable to these markers. Indeed, we were able to identify (Figure 4) a strong correlation between sMAdCAM levels and peak association rate of responses against the viral neutralization target, RBD in convalescent individuals. Also, we observed a relatively weaker but suggestive association with fold changes (day 0 and day 7) in sMIL index with that of viral Spike protein binding in plasma of the follow-up cohort.

**Figure 4.**
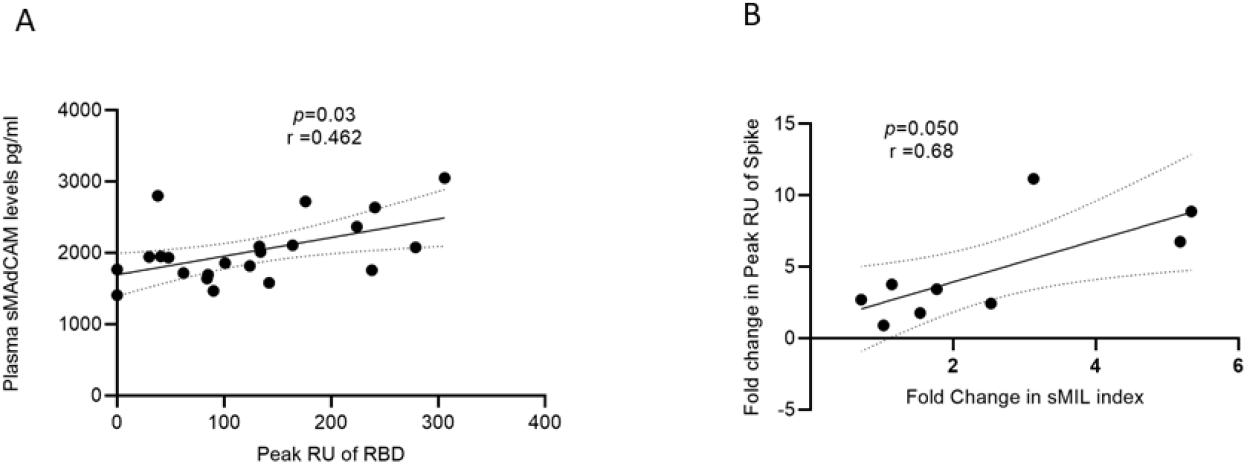
Association of sMAdCAM levels/sMIL index and virus specific binding in plasma(A) Association of plasma MAdCAM levels with Peak RU (association rate) of RBD binding among convalescent study participants. (B) Association of fold change between Peak RU for spike and sMIL index among study participants (n=9 out of 11) who were followed up. Fold change was calculated by dividing Day 7 values with those of Day 0. Fold change in Peak RU of 2 individuals could not be calculated as Day 0 values were undetectable (zero). Correlation analysis performed using non parametric Spearman Rank Correlation test. r indicates strength of correlation, p indicates significance.

## Discussion

Our study is the first to identify a putative integrated marker of inflammation and homeostatic immune migration that associates both with COVID-19 disease progression as well as generation of potentially neutralizing antibody (11) responses against SARS-CoV-2.

Considering the already demonstrated role of inflammatory cytokines in the development of pathology of COVID-19 (12) we undertook to evaluate how inflammatory profiles (Th1/Th2/Th17) correlated both cross-sectionally as well as longitudinally with varying disease states. Further, the role of an important emerging mediator of gut inflammation and mucosal homing, sMAdCAM, was examined in light of its significant role in the pathology of chronic inflammatory conditions including those with viral aetiology (13–17). This aspect of our study is especially noteworthy in the context of accumulating evidence for the gut as a potential sanctuary site for SARS-CoV-2 and persistent immunological sequelae observed in apparently virus free individuals (18–20). Whether altered sMAdCAM levels reflect changes in migration to various mucosal sites (gut to respiratory mucosa and vice versa) in response to SARS-CoV-2 viral infection remains to be established. Additionally, its role in microbial translocation, a well-established pathology for viruses such as HIV (21,22) and recently described in the context of SARS-CoV-2 infection needs to be evaluated (9).

Our in-patient cohort included participants at various stages of disease progression and included a group of 11 individuals where inflammatory profiles as well as anti-viral responses could be measured in a prospective manner relatively early after infection. These profiles were compared to a group of convalescent, disease free individuals that were presumed to have recovered from infection. We were also able to group study participants on the basis of anti-viral IgG and IgM responses which enabled a concurrent evaluation of inflammatory signatures with development of immune responses.

Our cytokine data highlights and confirms the predominance of IL-6 as an inflammatory mediator of pathology early following infection evinced through declining levels that trended inversely both with disease progression as well as transitioning of double negative (IgM-IgG) to IgG single positive anti-viral responses. Together with TNF, it was interesting to note that similar dysregulated levels of these cytokines were observed in apparently ‘convalescent’ individuals that, in some cases, had been discharged following not just resolution of symptoms but also repeated negative RT-PCR diagnoses in respiratory swabs. These observations may prove useful in optimising screening procedures of donors for plasma therapy that are often drawn from a pool of convalescent individuals (23), that as of now, are simply screened for the ability to mount anti-viral antibodies without accounting for sequelae such as ongoing inflammatory cytokine dysregulation. Also, stark differences in circulating IL-6, but not TNF levels were observed in male participants who exhibited higher levels of this cytokine. This observation is consistent with reports of milder pathology and faster recuperation following infection in women (24–26). Further, the small size of our female cohort with heterogenous age (implying both pre and post-menopausal status) limited our ability to delineate a role for sex hormones in the relatively lower inflammatory signature observed in women enrolled in our study. However, it is interesting to note that a very tight grouping for IL-6 levels was observed in these women, irrespective of their age (range, 20-72 years; Median, 38 years).

Our results with sMAdCAM are interesting, in light of the limited data available from select gut associated inflammatory pathologies, (27–30) however, when stratified by disease state (in-patient or convalescent status) as well as by gender an intriguing trend for this marker was observed. Both convalescent individuals and females had significantly elevated levels of sMAdCAM and conversely lower levels of IL-6. This observation led us to investigate whether the relationship between these 2 markers reflected an inflammatory/homeostatic dichotomy where favourable disease progression or convalescence was characterized by relative levels (or ratios) of these mediators. Indeed, our analysis, heretofore unreported, confirmed that only the ratio (but not levels in isolation) of sMAdCAM/IL-6 (sMIL index) was able to stratify infected individuals into varying disease states cross-sectionally, longitudinally, by gender and most importantly by immune competence as measured by class switched IgG anti-viral responses. Whether the sMIL index would serve as a similar correlate in other diseases involving inflammation and mucosal homeostasis remains to be evaluated.

The apparent link between sMIL index and class switched antibodies detected in advanced disease progression and convalescent individuals drove us to characterize plasma borne anti-viral humoral responses in our cohort against multiple viral targets NC, Spike and RBD, that are significant for diagnosis and vaccine development (31). Real-time binding profiles, through surface plasmon resonance (SPR), offered a more realistic recapitulation of in-vivo binding and were thus generated directly by flowing plasma over immobilized viral proteins. This analysis first revealed a good concordance between detected peak association rates (RU) of anti-NC responses and CLIA IgG indices obtained for the same target. Of note however is the observation that the SPR assay was able to detect binding against NC even in individuals that had undetectable levels as measured by CLIA, highlighting its possible utility as an early sero-diagnostic tool (32,33).

When comparing peak association rates (RUs) across disease progression (longitudinally) or disease states (in-patients and convalescents) responses to all 3 targets were clearly detectable by day 7 following diagnosis of infection. Also, convalescents uniformly had lower levels of these antibodies in circulation in keeping with their presumed viral clearance (34). Overall, in concordance with previous reports for both SARS and SARS-CoV-2 using endpoint assays (35,36), levels of anti-NC binding were detected early following infection, at high levels and appeared to be more persistent than those of anti-Spike responses. When stratified into groups reflecting maturation status of antibody responses (IgM/IgG status) we noticed gratifyingly that the relatively lower and less persistent anti-Spike responses could be explained by the observation that both anti-NC and RBD binding responses, but not anti-Spike responses were significantly observed at the IgM+IgG+ (double positive) stage suggesting earlier and probably better memory cell generation for these viral targets. We propose further exploration of these mechanisms may also help explain the observed rapid loss or waning to undetectable levels of these antibodies in some convalescent individuals (37,38) and also reported for SARS infection (39).

Finally, the observed correlation between sMAdCAM and sMIL index with peak anti-RBD RUs and fold change in anti-Spike responses respectively further confirm the association of apparently disparate arms of the immune response in generating anti-viral immunity and highlight the importance of inflammatory/mucosal homeostatic markers as putative positive immune correlates for efficacious vaccines (40–43).

## Limitations

The limitations of this study include a modest sized cohort that may reduce the impact of the observations and requires validation through larger prospective studies. Also, the follow-up data was based on 35 of 60 in-patients due to limited availability of sequential samples. Further, the date of diagnosis was considered as the onset of disease progression which may not have been the case in all cases.

## Conclusion

Our study is the first to report on the role for sMAdCAM in the pathology of and immunity against SARS-CoV-2 as well as on its utility together with IL-6 levels (sMIL index) in serving as an integrative disease progression marker for COVID-19.

## Data Availability

All data available for review upon request to corresponding authors.

## Author Contribution

## Acknowledgement

We are extremely grateful to Director General, Indian Council of Medical Research (ICMR), Ministry for Health & Family Welfare, Government of India for encouraging and supporting with resources, the pursuit of important research questions relevant to COVID-19 pathogenesis. Also, none of this work would have been possible without the active participation and support of participants to whom we are grateful.

## Notes

### Competing Interest Statement

The authors have declared no competing interest.

### Funding Statement

ICMR-NIRRH Intramural Core Support

### Author Declarations

KASTURBA HOSPITAL FOR INFECTIOUS DISEASES Institutional Review Board, Approval dated April 17,2020. ICMR-NIRRH Ethics Committee for Clinical Studies, Approval dated April 16, 2020; project no. 399/2020

