## Supplementary Information for "sMAdCAM:IL-6 ratio influences disease progression and anti-viral responses in SARS-CoV-2 infection"

Index

1. Supplemental Table 1. Study cohort characteristics
2. Supplemental Figure 1: Distribution of plasma inflammatory markers across various serological groups of SARS-CoV-2 infected individuals.
3. Supplemental Figure 2: Association of IL-6 with other cytokines and Ct for RT-PCR of different SARS-CoV-2 gene.
4. Supplemental Figure 3: Association of TNF with other cytokines and Ct for RT-PCR of different SARS-CoV-2 gene.
5. Supplemental Figure 4: Alterations in plasma inflammatory markers of SARS-CoV-2 infected individuals upon follow-up.
6. Supplemental Figure 5: Analysis of circulating lipopolysaccharides (LPS) with SARS-CoV-2 disease progression
7. Supplemental Figure 6: Comparison of plasma inflammation levels between male and female infected with SARS-CoV-2
8. Supplemental Figure 7: Association of IL-6 with days since SARS-CoV-2 infection and soluble MAdCAM
9. Supplemental Figure 8: Dynamic changes in CLIA index and Peak RU (association rates) of SARS-CoV-2 infected individuals

10.Supplemental Figure 9: Dynamic changes Peak RU (association rate) of SARS-CoV-2 infected individuals and Convalescent individuals

Supplemental Table 1. Study cohort characteristics

| Sample Id | Age (Yrs) | Gender (M/F) | Ct Value | | | Days post PCR test until sampling | (IgM) | (IgG) | CLIA IgG (Value) | Co-morbidities | Medication |
| --- | --- | --- | --- | --- | --- | --- | --- | --- | --- | --- | --- |
|  |  |  | N | O | S/R |  |  |  |  |  |  |
| U1 | 50s | M | 32 | 32 | 33 | 3 | + | + | 3.16 | HIV+ | Cefixime, Pan 40, Pcm, MVBC, Vit C (500mg), cough syrup |
| U2 | 50s | M | 19 | 21 | 20 | 9 | - | - | 1.55 | None | PCM 500, Pan 40, Levocet 5MG |
| U3 | 30s | M | 25 | 26 | 25 | 0 | - | - | - | TRF not traceable | NA |
| ^a^U4 | 30s | F | 31 | 29 | 28 | 11 | - | + | 5.94 | Diabetes mellitus | DD, MUBC, Metformin 500mg, Insulin , Pantop 40mg |
| U5 | 30s | M | 33 | 32 | 34 | 2 | + | + | 4.72 | Diabetes | Azithromycin,HCQ, Tamiflu, Vit C, Augmentin, |
| U6 | 70s | F | 28 | 25 | 26 | 11 | + | + | 5.28 | Diabetes, Hypertension | Azec, Pan40, Glimi, MVBC, Amlodipin, Inj PJ |
| U7 | 30s | F | 33 | 32 | 33 | 3 | + | + | 6.9 | None | NA |
| U8 | 40s | F | - | - | - | 17 | + | + | 2.6 | None | Pan , MVBC |
| ^a^U9 | 70s | M | 34 | 32 | 32 | 5 | - | - | 7.19 | None | Not available |
| U10 | 60s | M | 25 | 26 | 25 | 2 | - | - | 1.57 | Diabetes | HCQ, Tamiflu, Vit C, Pan 40, MVBC,Paracetamol |
| U11 | 60s | F | 33 | 32 | 33 | 3 | - | + | 8.23 | Diabetes mellitus | Azec, MVBC, Pan 40 |
| ^a^U12 | 30s | F | 37 | 33 | 32 | 11 | - | - | 5.85 | Diabetes mellitus | DD, MUBC, Pantop 40, Tenegliptin, Metformin |
| ^b^U13 | 50s | M | 26 | 24 | 25 | 6 | - | + | 6.47 | None | Augmentin, Para, Pan40, Vit C, MVBC |
| ^c^U14 | 50s | M | 35 | 28 | 28 | 6 | - | - | 5.47 | Diabetes mellitus | NA |
| ^a^U15 | 50s | M | 32 | 30 | 30 | 10 | - | - | - | Hypertension, Hyperlipidemia | Azec, MVBC, Pan 40 |
| ^a^U16 | 60s | F | 32 | 31 | 32 | 6 | - | - | 8.8 | None | MVBC, Pan 40 |
| U17 | 60s | M | - | 32 | - | 19 | - | + | 7.83 | Diabetes, Hypertension | Telma 40 mg, amlo 5mg, rosuvastatin 5mg, levocet10mg, MVBC, Pan 40, PCM 500mg, Teneligliptin/metformin 20/500 |
| U18 | 40s | M | 27 | 26 | 26 | 11 | - | + | 7.08 | None | PCM, Pan 40, MVBC, Azec |
| ^a^U19 | 50s | M | 23 | 22 | 22 | 9 | - | + | 3.36 | Diabetes, Immune thrombocytopenia | Azec 500, mvbc, pan 40, pcm 500 |
| U20 | 40s | M | 30 | 30 | 30 | 3 | - | - | - | None | NA |
| U21 | 40s | M | 24 | 22 | 23 | 5 | - | + | 4.07 | None | Augmentin, Calpol 650, MVBC, Vit C, Pan 40 |
| ^a^U22 | 30s | M | 31 | 31 | 31 | 3 | - | + | 5.93 | None | Azec, Pan 40, MVBC |
| U23 | 40s | M | 32 | 31 | 31 | 3 | - | + | 7.18 | Diabetes | Azec, Metrogyl, Vizyla |
| U24 | 50s | M | 28 | 26 | 27 | 5 | - | - | - | Diabetes, Hypertension, Stroke | Augmentin, sinarest, calpol, MVBC, Deplatt A, Zeryl, Atorva, Glimy M2, Felicita OD, Pan 40 |
| ^b^U25 | 20s | M | 20 | 23 | 19 | 11 | - | + | 7.37 | None | PCM, Cough syrup, Pantaprazole |
| S10 | 40s | M | ₋ | ₋ | 32 | 0 | - | - | - | Hypertension, Heart disease | Azec 500 mg, Pan 40, MVBC, PCM 500 mg, Vit C, Vit A |
|  |  |  |  |  | Neg | 7 | + | + | 4.23 |  |  |
| S11 | 50s | M | ₋ | ₋ | 21 | 0 | - | - | 3.98 | Diabetes mellitus | PCM 500, Pan 40, Azee 500, MVBC |
|  |  |  |  |  | NA | 7 | + | + | 9.69 |  |  |
| S20 | 70s | M | ₋ | ₋ | 26 | 0 | - | - | - | Diabetes, Hypertension | Pan, Vit C, Azec, MVBC |
|  |  |  |  |  | Neg | 7 | - | - | 7.2 |  |  |
| S21 | 40s | M | ₋ | ₋ | NA | 0 | - | - | - | None | Azec, PCM, Pan, ondem, MVBC, Vit C, Zinc |
|  |  |  |  |  | NA | 7 | + | + | 6.37 |  |  |
| S23 | 40s | M | ₋ | ₋ | 28 | 0 | - | - | 4.02 | Diabetes, Dyslipidemia | Azithromycin, Tamiflu, for diabetes: Rozavel, Zita plus, Glimisave |
|  |  |  |  |  | NA | 7 | - | + | 9.19 |  |  |
| S24 | 50s | M | ₋ | ₋ | 24 | 0 | - | - | - | None | Azec 500, paracetamol 500, Pau 40, Ewseb 4mg, MUBC, Vit C, Zinc, cough syrup |
|  |  |  |  |  | 33 | 7 | - | - | 3.26 |  |  |
| S25 | 40s | M | ₋ | ₋ | 24 | 0 | + | + | 3.92 | None | Azec, PCM, Emset, MVBC, vit C, zinc |
|  |  |  |  |  | NA | 7 | + | + | 8.03 |  |  |
| S26 | 20s | M | ₋ | ₋ | 33 | 0 | - | - | - | None | NA |
|  |  |  |  |  | Neg | 7 | - | + | 1.74 |  |  |
| S27 | 30s | M | ₋ | ₋ | 37 | 0 | + | + | 7.79 | Depression | NA |
|  |  |  |  |  | Neg | 7 | - | + | 7.64 |  |  |
| S29 | 30s | M | ₋ | ₋ | 25 | 0 | - | - | 3.42 | None | NA |
|  |  |  |  |  | Neg | 7 | - | - | 5.18 |  |  |
| S30 | 40s | F | ₋ | ₋ | NA | 0 | - | - | - | Hypertension, Hypothyroidism | Azee 500, Pan40, MVBC, PCM 500, Thyronorm 100ug, continue antihypertensive, Cough syrup, HCQ, Vit C, Vit A |
|  |  |  |  |  | NA | 7 | - | + | 6.15 |  |  |
| S122 | 70s | M | ₋ | ₋ | 32 | 0 | - | - | NA | None | NA |
|  |  |  |  |  |  | 7 | - | + | NA |  |  |
| S123 | 40s | M | ₋ | ₋ | 32 | 0 | - | - | NA | None | NA |
|  |  |  |  |  |  | 7 | - | + | NA |  |  |
| S125 | 60s | F | ₋ | ₋ | 28 | 0 | - | - | NA | None | NA |
|  |  |  |  |  |  | 7 | - | + | NA |  |  |
| S132 | 40s | M | ₋ | ₋ | 30 | 0 | + | + | NA | None | NA |
|  |  |  |  |  |  | 7 | + | + | NA |  |  |
| S133 | 60s | M | ₋ | ₋ | 24 | 0 | - | - | NA | Diabetes,Hypertension,Asthama | NA |
|  |  |  |  |  |  | 7 | - | + | NA |  |  |
| S171 | 30s | F | ₋ | ₋ | 29 | 0 | - | + | NA | None | NA |
|  |  |  |  |  |  | 7 | + | + | NA |  |  |
| S172 | 40s | F | ₋ | ₋ | 38 | 0 | + | + | NA | Thyroid | NA |
|  |  |  |  |  |  | 7 | + | + | NA |  |  |
| S173 | 30s | M | ₋ | ₋ | 29 | 0 | - | - | NA | None | NA |
|  |  |  |  |  |  | 7 | + | + | NA |  |  |
| S174 | 40s | M | ₋ | ₋ | 24 | 0 | - | + | NA | None | NA |
|  |  |  |  |  |  | 7 | - | + | NA |  |  |
| S175 | 70s | M | ₋ | ₋ | 24 | 0 | - | + | NA | Hypertension | NA |
|  |  |  |  |  |  | 7 | + | + | NA |  |  |
| S177 | 60s | M | ₋ | ₋ | 30 | 0 | + | + | NA | Diabetes,Hypertension | Mezopenum 1g, Pan 40mg, MPS 500mg, Clexane 0.4cc |
|  |  |  |  |  |  | 7 | + | + | NA |  |  |
| S178 | 50s | M | 22 | 24 | ₋ | 0 | NA | NA | NA | None | NA |
|  |  |  |  |  |  | 7 | NA | NA | NA |  |  |
| S179 | 40s | M | 38 | 38 | ₋ | 0 | - | - | NA | None | NA |
|  |  |  |  |  |  | 7 | + | + | NA |  |  |
| S183 | 20s | M | 32 | 34 | ₋ | 0 | - | - | NA | None | NA |
|  |  |  |  |  |  | 7 | - | + | NA |  |  |
| S185 | 70s | F | 34 | 34 | ₋ | 0 | + | + | NA | None | NA |
|  |  |  |  |  |  | 7 | + | + | NA |  |  |
| S187 | 50s | F | 38 | 38 | ₋ | 0 | - | - | NA | None | NA |
|  |  |  |  |  |  | 7 | + | - | NA |  |  |
| S189 | 30s | M | 37 | 37 | ₋ | 0 | - | - | NA | Hypertension | NA |
|  |  |  |  |  |  | 7 | - | - | NA |  |  |
| S192 | 50s | M | ₋ | 39 | ₋ | 0 | - | - | NA | None | NA |
|  |  |  |  |  |  | 7 | - | + | NA |  |  |
| S193 | 40s | M | 33 | 33 | ₋ | 0 | + | + | NA | None | NA |
|  |  |  |  |  |  | 7 | + | + | NA |  |  |
| S195 | 50s | M | 25 | 24 | ₋ | 0 | - | - | NA | Diabetes, Heart disease | NA |
|  |  |  |  |  |  | 7 | - | + | NA |  |  |
| S196 | 60s | F | 18 | 18 | ₋ | 0 | - | - | NA | Diabetes,Hypertension | NA |
|  |  |  |  |  |  | 7 | - | - | NA |  |  |
| S197 | 30s | F | 37 | ₋ | ₋ | 0 | - | - | NA | None | NA |
|  |  |  |  |  |  | 7 | - | - | NA |  |  |
| S198 | 40s | M | 38 | 37 | ₋ | 0 | + | - | NA | None | NA |
|  |  |  |  |  |  | 7 | + | - | NA |  |  |
| S199 | 30s | M | 32 | 34 | ₋ | 0 | - | + | NA | None | NA |
|  |  |  |  |  |  | 7 | - | + | NA |  |  |
| C26 | 40s | M | ₋ | 35 | ₋ | 32 | - | + | Plasma NA | None | NA |
| C28 | 40s | M | ₋ | 34 | ₋ | 35 | - | + | Plasma NA | Diabetes, Hypertension, Heart disease | NA |
| C29 | 30s | M | ₋ | 29 | ₋ | 40 | - | + | Plasma NA | None | NA |
| C30 | 20s | M | ₋ | 36 | ₋ | 35 | - | + | 2.6 | None | NA |
| C31 | 30s | M | ₋ | 37 | ₋ | 30 | - | + | 7.78 | COPD, Diabetes | NA |
| C32 | 20s | F | ₋ | ₋ | ₋ | 19 | - | + | 4.15 | NA | NA |
| C33 | 50s | M | ₋ | ₋ | ₋ | 19 | - | + | 4.75 | NA | NA |
| C42 | 20s | M | ₋ | ₋ | ₋ | 33 | - | - | - | NA | NA |
| C43 | 20s | M | ₋ | 25 | ₋ | 39 | - | + | 3.75 | NA | NA |
| C44 | 40s | M |  |  |  | NA | - | + | 4.48 | NA | NA |
| C45 | 20s | M | - | 24 | 26 | 37 | - | + | 2.63 | NA | NA |
| C46 | 40s | M | - | + | - | 31 | + | + | 5.31 | NA | NA |
| C47 | 20s | M | - | 27 | - | 41 | - | + | 4.89 | NA | NA |
| C48 | 30s | M |  |  |  | NA | - | - | - | NA | NA |
| C49 | 40s | M | + | + | - | 60 | - | - | - | NA | NA |
| C50 | 20s | F | - | 37 | - | 62 | - | + | 2.92 | NA | NA |
| C51 | 30s | M | - | - | 28 | 57 | - | + | 6.57 | NA | NA |
| C52 | 20s | M | - | - | - | 48 | + | + | - | NA | NA |
| C53 | 50s | M | - | 23 | - | 38 | - | + | 2.58 | NA | NA |
| C54 | 20s | F | - | 33 | - | NA | - | + | - | NA | NA |
| C55 | 30s | M | NA | NA | NA | 36 | - | + | - | NA | NA |
| C56 | 30s | M | - | 30 | - | 40 | - | + | 9.47 | NA | NA |
| C57 | 20s | F | NA | NA | NA | 52 | - | + | NA | None | None |
| C58 | 30s | F | NA | NA | NA | 55 | - | + | NA | None | None |

Footnote: U: 1st set of 25 in-patients: (U1 – U25); C: Convalescent plasma candidates (C26 –C58); S: In- patients recruited and followed up (Day 0 and Day 7, S10 – S199); NA – Not Available; NT – Not Tested; a, Asymptomatic individuals; b, Individuals transferred to other hospitals; c, Discharge information not available. All others were discharged after two consecutive negative PCR results or resolution of symptoms. Co-morbidities where documented are listed. Rapid Antibody test had 77% concordance with CLIA.

Supplemental Figure 1: Distribution of plasma inflammatory markers across various serological groups of SARS-CoV-2 infected individuals. Comparison of plasma levels of (A) IL-6 in 19 healthy, seronegative controls, in-patients - IgG-/IgM- (n=22), IgG+/IgM+ (n=11), IgG+/IgM- (n=14) and sero-convalescent (n=24) groups (B) TNF in 3 healthy, seronegative controls, in-patients - IgG-/IgM- (n=22), IgG+/IgM+ (n=11), IgG+/IgM- (n=14) and sero-convalescent (n=22) groups and (C) Soluble MAdCAM concentration in 19 healthy, seronegative controls, in-patients - IgG-/IgM- (n=38), IgG-/IgM+ (n=13) IgG+/IgM+ (n=25), IgG+/IgM- (n=17) and sero-convalescent (n=24) groups. Red coloured symbols represent asymptomatic individuals. Pink coloured symbols indicate IgG-/IgM- individuals in sero-convalescent group. Statistical significance was calculated by unpaired Mann- Whitney U-test; *, p < 0.05; **, p < 0.01; ***, p<0.001 and ****, p<0.0001

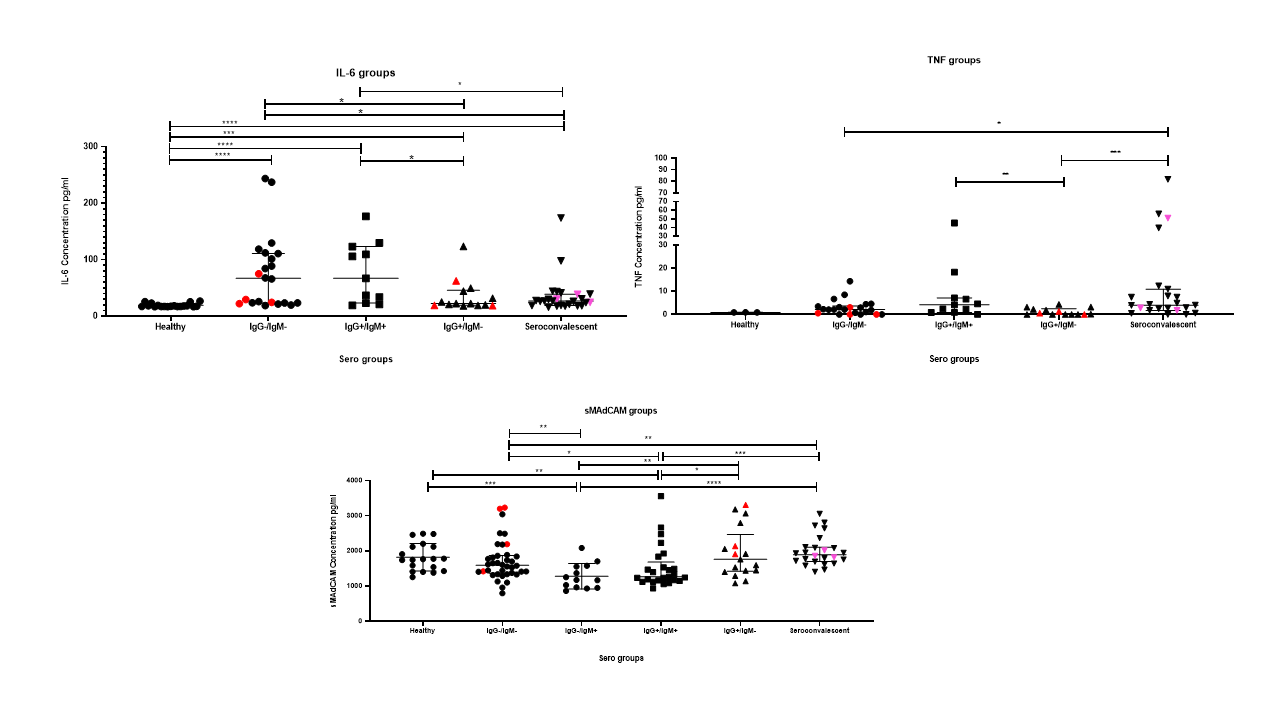

Supplemental Figure 2: Association of IL-6 with other cytokines and Ct for RT-PCR of different SARS-CoV- 2 gene. Association of plasma IL-6 levels with cytokines (n=69) (A) IL-2, (B) IL-4, (C) IL-10, (D) IFNγ, (E) TNF, and (F) IL-17A as well as with Ct values for viral gene targets (G) N (n=23); (H) O (n=36) and (I) S (n=50) as evaluated by real time PCR. Undetectable levels of cytokines have been assigned values of 0.

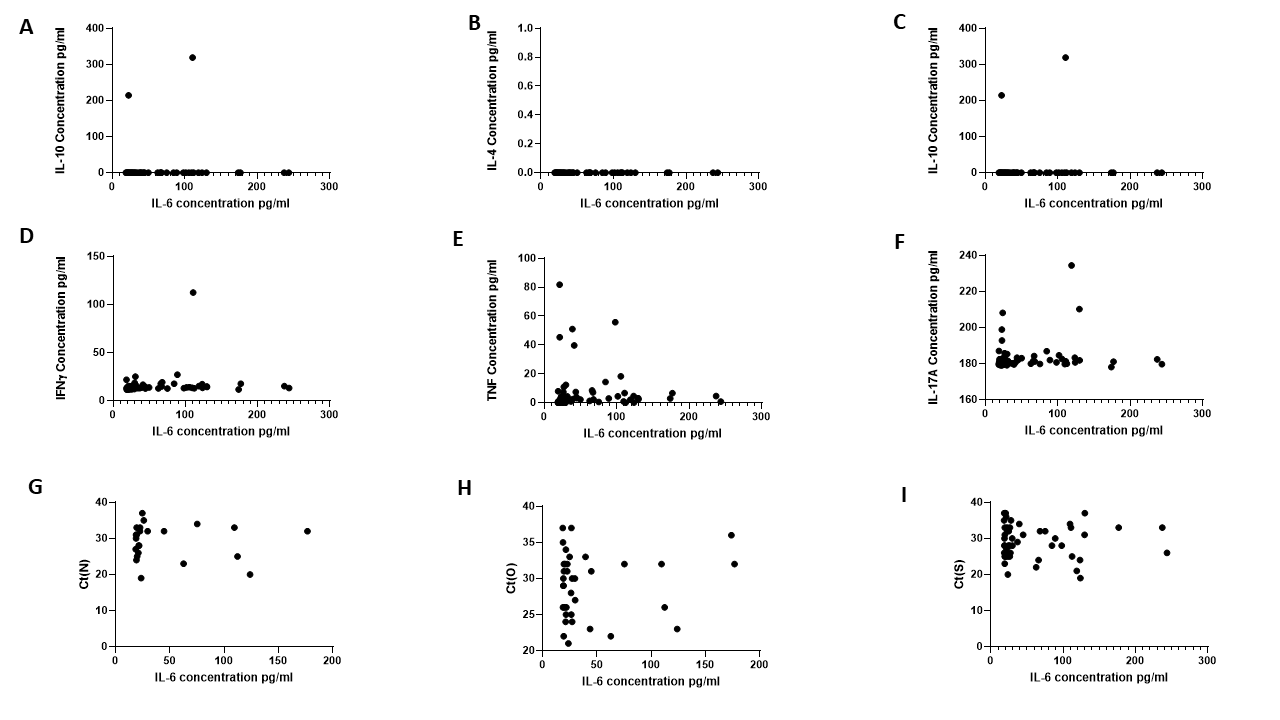

Supplemental Figure 3: Association of TNF with other cytokines and Ct for RT-PCR of different SARS- CoV-2 gene. Association of plasma TNF levels with cytokines (n=69) (A) IL-2, (B) IL-4, (C) IL-10, (D) IFNγ and (E) IL-17A as well as with Ct values for viral gene targets (F) N (n=23); (G) O (n=36) and (H) S (n=50) as evaluated by real time PCR. Undetectable levels of cytokines have been assigned values of 0.

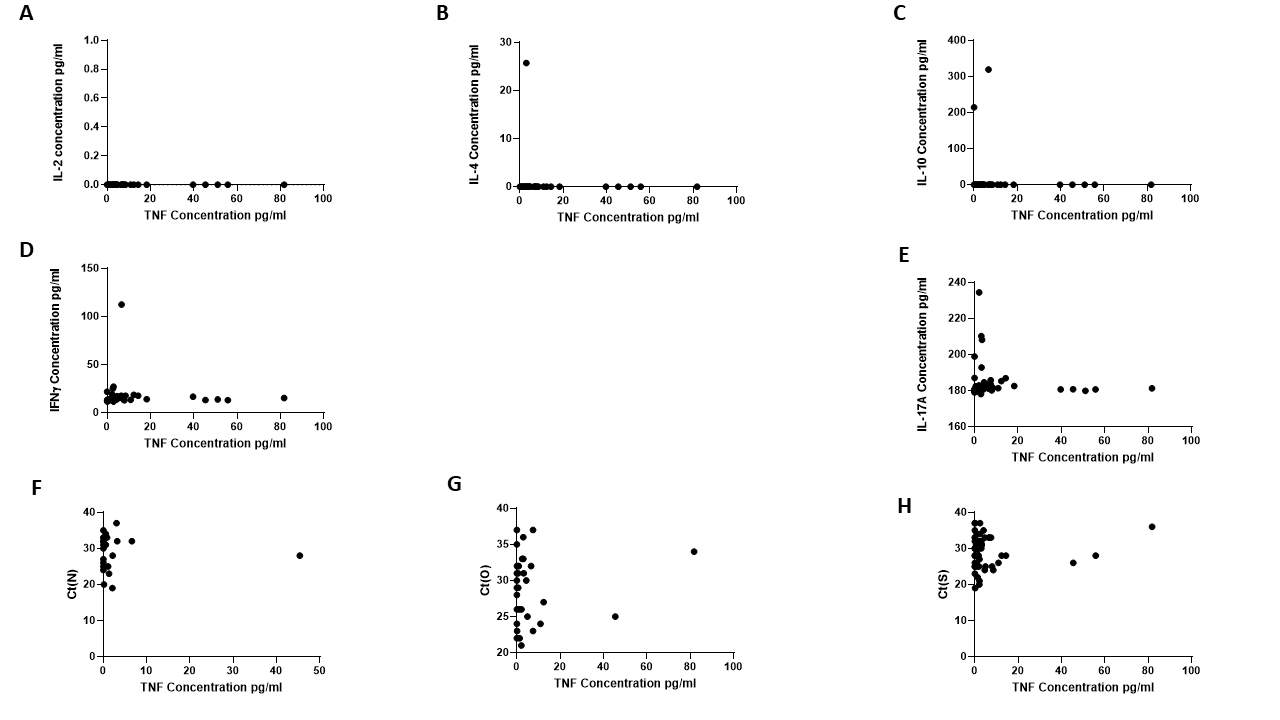

Supplemental Figure 4: Alterations in plasma inflammatory markers of SARS-CoV-2 infected individuals upon follow-up. Change in plasma levels of (A) IL-6(n=11) (B) TNF(n=11) and (C) Soluble MAdCAM (n=35) between day 0 and day 7. Statistical significance was calculated by Wilcoxon matched-pairs signed rank test.

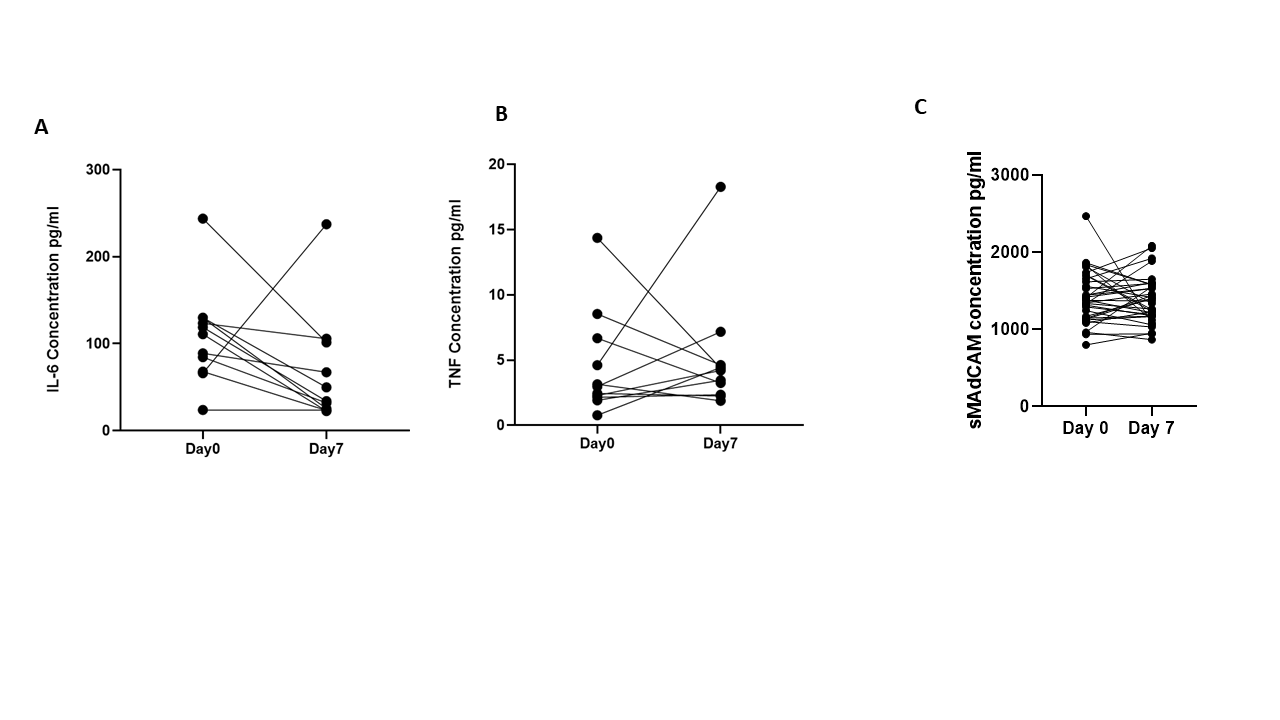

Supplemental Figure 5: Analysis of circulating lipopolysaccharides (LPS) with SARS-CoV-2 disease progression A. Comparison of LPS between IgG-/IgM- (n=10), IgG+/IgM+ (n=5), IgG+/IgM- (n=10) and convalescent (n=4) groups. B. Variation in LPS among in-patients (n=25) and in convalescent (n=4) C. Differences in LPS between male (n=21) and female (n=8). D-E. Association of circulating LPS with (D) IL-6 and (E) sMAdCAM (n=29). Red coloured symbols represent asymptomatic individuals. Statistical analysis was performed using Graphpad Prism 8.0 Mann-Whitney U-test was used to compare unpaired groups. *, p < 0.05; **, p < 0.01, ***, p<0.001; and ****, p<0.001. Correlation analysis was performed using non parametric Spearman Rank Correlation test.

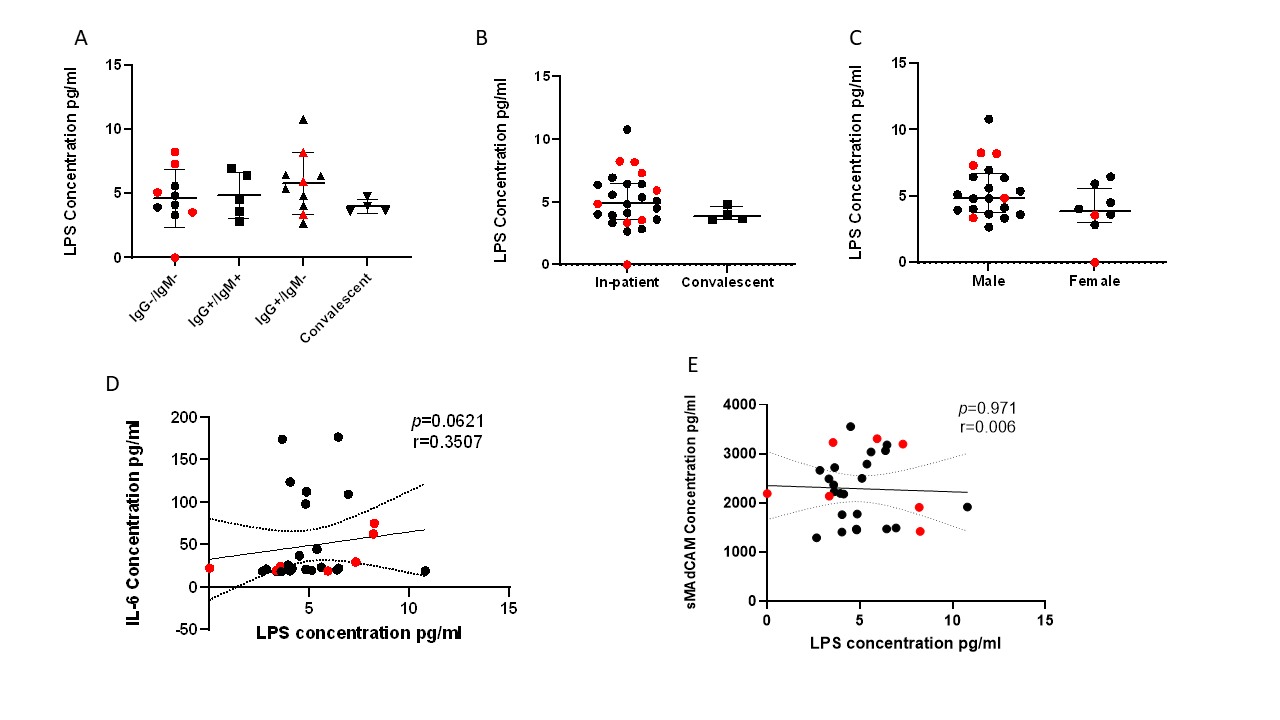

Supplemental Figure 6: Comparison of plasma inflammation levels between male and female infected with SARS-CoV-2. Differences in plasma levels of (A) IL-6 (B) TNF and (C) Soluble MAdCAM between male (n=18) and female (n=7). Red coloured symbols represent asymptomatic individuals. Statistical significance was calculated by unpaired Mann-Whitney U-test; *, p < 0.05; **, p < 0.01; and ***, p<0.001.

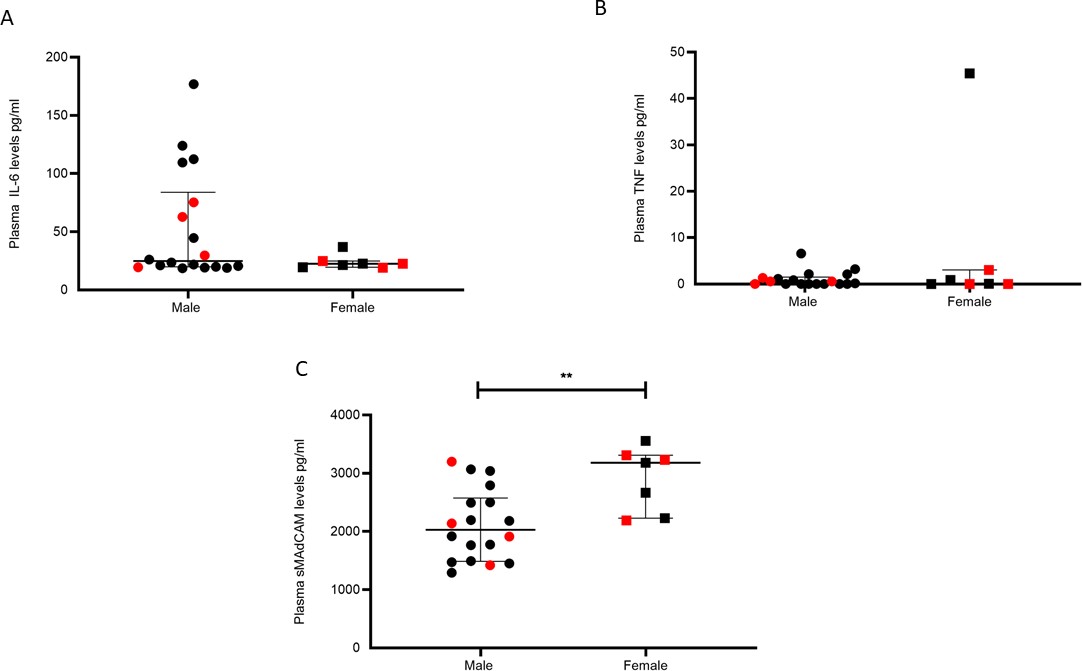

Supplemental Figure 7: Association of IL-6 and soluble MAdCAM with days since SARS-CoV-2 infection. A-B. Association of (A) IL-6 levels (n=47) and (B) soluble MAdCAM levels(n=95) in plasma of SARS-CoV-2 infected study participants with days since SARS-CoV-2 confirmation by PCR. (C) Association of IL-6 and soluble MAdCAM among in-patients (n=47) *r* indicates strength of correlation, *p* indicates significance. Negative r value indicates inverse correlation. Red coloured symbols represent asymptomatic individuals. Statistical significance was calculated by Spearman correlation analysis

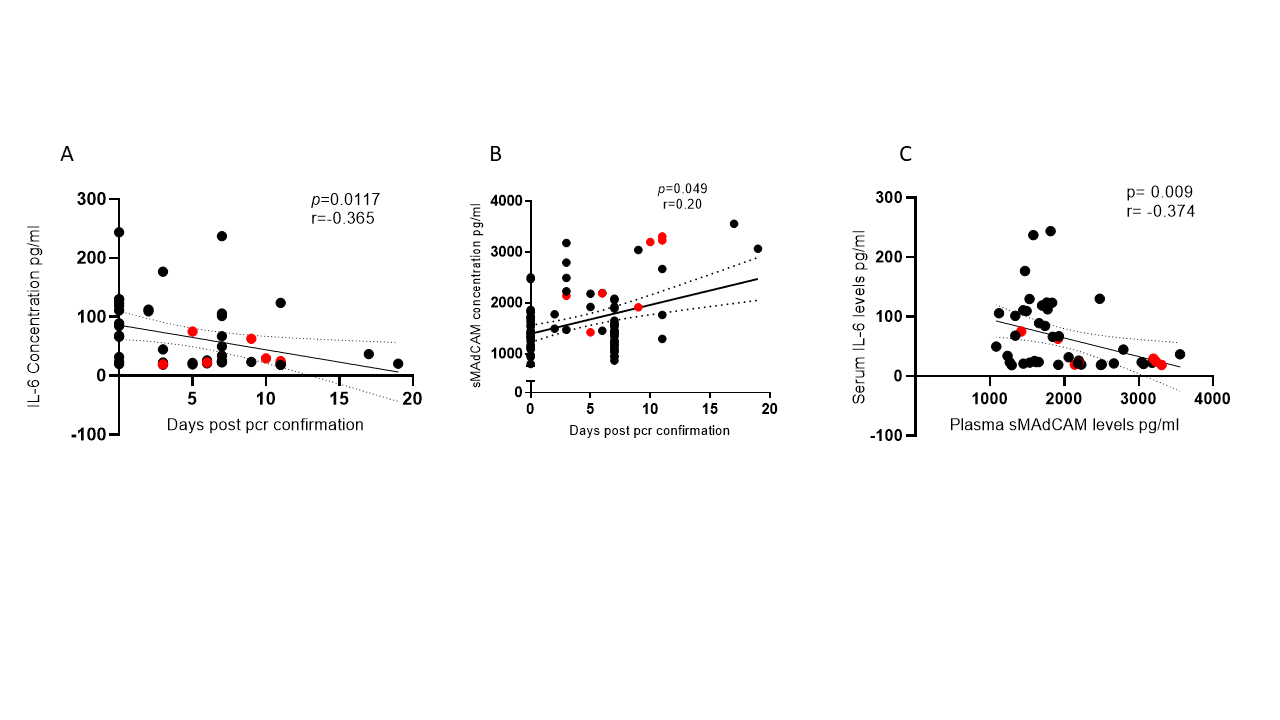

Supplemental Figure 8: Dynamic changes in CLIA index and Peak RU (association rates) of SARS- CoV-2 infected individuals. Variation in levels of (A) CLIA (NC specific) index (B) peak RU for Nucleocapsid, (C) Spike and (D) RBD among study participants followed up in this study. Statistical significance was calculated by Wilcoxon matched-pairs signed rank test; *, p < 0.05; **, p < 0.01; and ***, p<0.001.

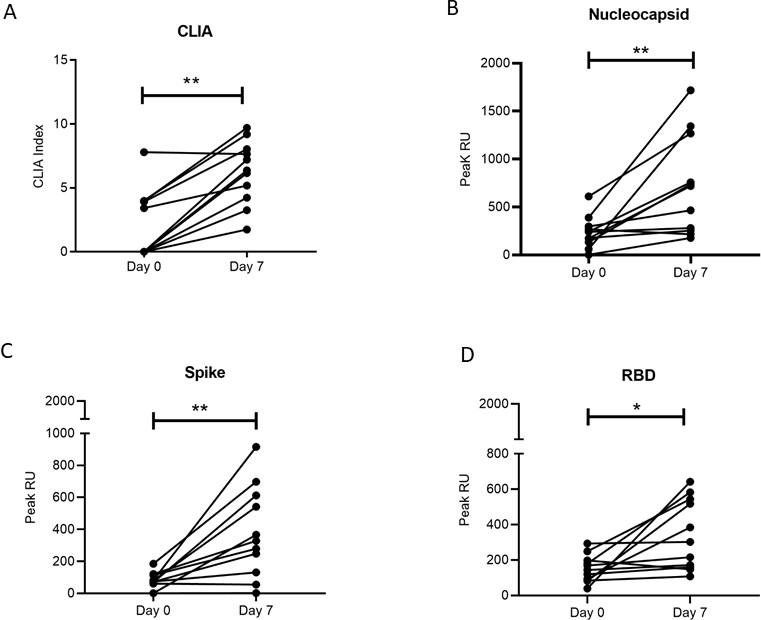

Supplemental Figure 9: Dynamic changes Peak RU (association rate) of SARS-CoV-2 seronegative (n=3), infected individuals and Convalescent individuals. Variation in peak RU for (A) Spike, (B) RBD and (C) Nucleocapsid, compared at day 0 (n=11), day 7 (n=11) and in convalescent (n=22) group. Pink coloured symbols indicate IgG-/IgM- individuals in convalescent group. Statistical significance was calculated by Wilcoxon matched-pairs signed rank test and Mann-Whitney U- test; *, p < 0.05; **, p < 0.01; and ***, p<0.001.

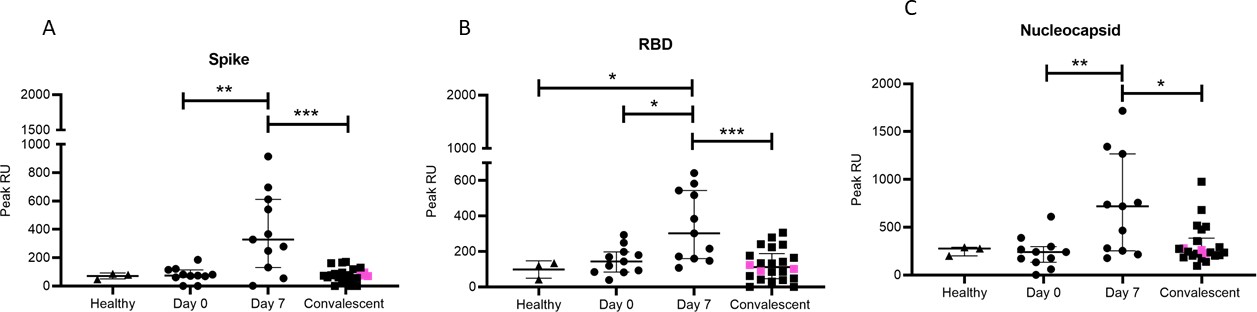
